# Reliance on Prior Expectations in Psychosis: A Systematic Review and Meta-Analysis of Perceptual Tasks

**DOI:** 10.64898/2026.03.31.26349835

**Authors:** Chantal Miller-Silva, Benjamin JG Illingworth, Kirsten Martey, Tamara Mujirishvili, Franciska de Beer, Dan Siskind, Graham K Murray

**Author notes:** **Conflicts of Interest** All authors declare no conflicts of interest. **Author Contributions** C Miller-Silva: Conceptualisation, data curation, investigation, formal analysis, funding acquisition, writing (original draft; review & editing), project administration, software, investigation, methodology, validation, visualisation. BJG Illingworth: Formal analysis, writing (review & editing), validation. K Martey: Data curation, investigation, validation. T Mujirishvili: Conceptualisation, writing (review & editing), methodology. F de Beer: Conceptualisation, writing (review & editing), methodology. D Siskind: Conceptualisation, writing (review & editing), methodology. GK Murray: Conceptualisation, investigation, formal analysis, funding acquisition, writing (review & editing), supervision, project administration, resources, methodology, validation.

## Abstract

**Background:** The highly influential predictive processing theory of psychosis posits that symptoms arise from imbalances in the weighting of predictions (*priors*) and sensory evidence. Despite this theory’s increasing prominence, studies often present conflicting results. This is particularly problematic as findings from single tasks with modest sample sizes are frequently used to advance a theory for a generalised altered reliance on priors in psychosis.

**Methods:** This study presents a random-effects, multi-level meta-analysis (PROSPERO CRD42024574379) evaluating evidence for aberrant reliance on priors in psychosis across perceptual tasks. The search identified articles in Embase, MEDLINE, APA PsycINFO, and APA PsycArticles published between 1^st^ January 2005 and 31^st^ October 2024, with risk of bias assessed using the Newcastle-Ottawa Scale. Included articles (34 results from 27 studies) compared adults with schizophrenia-spectrum psychosis (SZ; *n* = 904) to healthy controls (*n* = 1,039) on behavioural measures representing reliance on priors.

**Results:** Results provided no evidence for atypical reliance on priors in psychosis (*g* =.03, 95% CI [-0.27, 0.34]; *p* =.818) or associations with delusions (6 results; SZ = 183; *r* = -.16, 95% CI [-0.51, 0.19]; *p* =.293) or hallucinations (10 results; SZ = 370; *r* =.04, 95% CI [-0.28, 0.36]; *p* =.780). In contrast with the theory that psychosis may differentially affect priors at different levels of the cognitive hierarchy, a sub-group analysis indicated that a two-level hierarchical model of priors did not account for conflicting results (*F*(1,32) = 0.1, *p* =.758).

**Conclusion:** These findings do not suggest that psychosis is associated with a generalised predictive processing deficit spanning multiple aspects of perception.

## Introduction

Despite schizophrenia-spectrum psychosis affecting 23.6 million people worldwide (Solmi et al., 2023), conclusive evidence for an underlying cognitive mechanism of illness is yet to be identified. One proposal cites imbalances in predictive processing: the integration of sensory evidence with prior expectations (*priors*; Fletcher & Frith, 2009). While predictive mechanisms enable efficient perception, reliance on expectations can cause non-veridical perception even in health. For example, changes in an individual’s reliance on priors, often termed *strength*, is a possible mechanism for numerous illusions (Hohwy, Roepstorff, & Friston, 2008; Kok, Bains, van Mourik, Norris, & de Lange, 2016). A body of literature has extended this discussion to address positive symptoms of psychosis: anomalous percepts (hallucinations) and beliefs (delusions; Sterzer et al., 2018). However, studies often employ a variety of different tasks with small samples and report conflicting results. Studies have reported both stronger (e.g., Powers, Mathys, & Corlett, 2017) and weaker (e.g., Schmack, Schnack, Priller, & Sterzer, 2015) priors in psychosis, while others suggest no significant difference compared to controls (Daalman, Verkooijen, Derks, Aleman, & Sommer, 2012). Likewise, studies disagree on which symptoms are underpinned by aberrant priors. Findings often demonstrate a relationship with one symptom; for example, Colbert, Peters, & Garety (2010) reported stronger priors related to delusional traits but not hallucinations. However, a comprehensive theory should explain common co-existent symptoms. One explanation for these discrepancies posits a hierarchy of priors reflecting the interlinked cortical hierarchy described in predictive processing (Hohwy, 2013; Rao & Ballard, 1999). In this account, weaker lower-level sensory priors trigger a compensatory strengthening of priors reflecting higher-order beliefs about the environment (Sterzer et al., 2018). Haarsma et al. (2020) probed this theory using separate tasks, finding weaker *perceptual* (low-level) priors alongside stronger *cognitive* (high-level) priors in first-episode psychosis (FEP). This theory not only has the potential to reconcile findings but also offers an account of co-existent symptoms, with weaker perceptual priors triggering anomalous perceptions and stronger cognitive priors underpinning delusions. However, the possibility of a differential reliance on distinct types of priors across the cortical hierarchy has not yet been tested across paradigms or with a large patient cohort.

The diversity of paradigms and results discussing reliance on priors in psychosis calls for a systematic analysis. The current study conducts a meta-analysis comparing prior strength in schizophrenia-spectrum psychosis versus healthy controls. This offers increased statistical power to assess whether an altered reliance on priors can explain psychotic symptoms across tasks. Subsequent analyses investigate whether differential effects can be explained by a hierarchy of priors, offering a much-needed empirical evaluation of this frequently-cited theory. As a theory of psychosis should explain symptoms, separate meta-analyses are conducted to address the relationship between reliance on priors and delusions and hallucinations. Overall, these analyses explore whether discussions of universally strong or weak priors in psychosis offer an oversimplified description of interdependent mechanisms that reflect a hierarchy of priors and explain co-existent symptoms. Overall, this contributes a systematic summary of quantitative results and suggests directions for future research.

## Methods

### Systematic Review Protocol

The review protocol (Miller-Silva, Murray, & Martey, 2024) was pre-registered in the PROSPERO registry (CRD42024574379) to reduce the risk of bias and provide a transparent record of decision-making. Analyses not featured in this protocol are labelled ‘exploratory’.

#### Population & Controls

This review consisted of studies with medicated or unmedicated schizophrenia-spectrum psychosis patients, including FEP, diagnosed by a clinician or researcher using a widely accepted diagnostic tool. Studies with participants classified as at-risk mental state (ARMS) or with psychosis caused by either a short-term, drug-induced state or neurological disorder were excluded. Controls had no known history of psychosis, ARMS status, or neurological disorder. All participants were adults (≥ 18 years).

#### Intervention

This review defines priors as beliefs about upcoming sensory information that can potentially bias perception. Included studies featured prior-inducing stimuli as the intervention, and these could take various forms (e.g., written information, stimulus conditioning). Only paradigms implementing a controlled manipulation of priors were included, meaning that reliance on priors was unequivocally tested. This criterion was necessary given that predictive processing is often cited as a unified theory of brain function (Clark, 2013) therefore many task outcomes could be interpreted as reflecting the use of predictive cues. This led to the exclusion of some commonly-used tasks that assess patients’ susceptibility to illusions (e.g., Kaliuzhna et al., 2019) as these paradigms often have no direct manipulation of priors but assume that task performance relies on prior knowledge of natural scene statistics. A systematic review of visual illusion tasks in patients was recently completed (Costa, Costa, Pessoa, Caixeta, & Maior, 2023).

#### Outcomes

Study results represented participants’ reliance on priors. Included data were quantitative, continuous, and behavioural (e.g., accuracy, response bias). Imaging results were excluded. Studies investigated the effect of priors on perception (e.g., visual detection, audiovisual integration, motion processing). Tasks with a main outcome assessing metacognition (confidence, agency) or other higher-level abilities (e.g., planning, learning, memory, mentalising, time perception, language, emotion recognition) were excluded. Representational momentum, time-to-collision, and jumping to conclusions tasks were thus excluded. Given the broad definition of ‘priors’ and variety of paradigms, this aimed to limit the scope of the review to a more similar, defined subset of perceptual tasks.

Data examining the relationship between prior strength and hallucinations and delusions in patients were also extracted in the form of group comparisons (e.g., patients with versus without hallucinations) or correlations with symptom scores. Results for psychosis-like experiences in controls (schizotypy) were not extracted.

#### Study Design

Quantitative English-language papers were included in the review. Reviews (including systematic reviews and meta-analyses) were excluded as they presented no original data. Case reports were excluded.

### Literature Search & Screening

The literature search was conducted between 7^th^ August 2024 and 31^st^ October 2024 using Embase (including a grey literature search of bioRxiv and medRxiv), MEDLINE, APA PsycINFO, and APA PsycArticles (full search strategy in Appendix A). The bibliographies of included articles were hand-searched for additional references. The search filtered articles from 1^st^ January 2005 onwards. This period covers the most relevant papers given that a seminal study on prediction and psychosis was published that year (Ford & Mathalon, 2005), followed by a review of predictive processing in schizophrenia (Fletcher & Frith, 2009).

This review followed the Preferred Reporting Items for Systematic Reviews and Meta-Analyses (PRISMA; Page et al., 2021; Appendix B). The screening process for articles was independently conducted in Rayyan (Ouzzani, Hammady, Fedorowicz, & Elmagarmid, 2016) by C.M-S and K.M.

### Data Extraction & Quality Assessment

Data extraction was conducted independently by C.M-S and K.M in Microsoft Excel, with conflicts reviewed by G.K.M. The number of participants and summary statistics for age and sex were extracted for patients and controls separately. For patients, the diagnosis; diagnostic method; chlorpromazine-equivalent dosage; and scores for the Positive and Negative Syndrome Scale (PANSS; Kay, Fiszbein, & Opler, 1987), Peters, Joseph, & Garety (1999) Delusion Inventory (PDI), and Cardiff Anomalous Perceptions Scale (CAPS; Bell, Halligan, & Ellis, 2006) were extracted. Exclusion criteria for both participant groups and covariates were noted for the quality assessment.

For each task, descriptions of the paradigm, prior, and outcome measure were recorded. The type of prior in each task was coded as ‘perceptual’ or ‘cognitive’. Perceptual priors were defined as inducing implicit and/or automatic lower-level constraints on sensory processing. Cognitive priors induced higher-level beliefs that provided contextual abstractions, involving rapidly learned schemas, semantics, or explicit probabilities. For example, the (false) perception of a sound following exposure to a conditioned visual stimulus is the effect of a perceptual prior: the belief that the sound and visual stimulus always co-occur. In contrast, explicit information about the likelihood of perceiving a stimulus represents a cognitive prior. C.M-S and K.M classified the priors, with conflicts resolved by G.K.M. Results were reported as group means and standard deviations, effect sizes, or correlation coefficients. If these statistics were missing from the article, an online plot digitiser (PlotDigitizer, 2025) was used to extract the group summary statistics from figures, or authors were contacted for the missing data. A qualitative summary of each finding and key interpretations by the authors were also recorded.

One anticipated challenge for this review was multiplicity as studies often compare multiple patient groups to controls. Pre-registered inclusion criteria sought to reduce multiplicity and the risk of bias associated with the subjective selection of results for inclusion. For example, some studies feature separate groups for patients with and without active symptoms. As the research question focuses on the effect of psychosis on prior strength, results for patients with and without active symptoms were combined and compared to controls. The relationship between priors and individual symptoms were then explored in later analyses. Results from replications of paradigms with the same participant groups were averaged.

C.M-S and K.M independently completed a quality assessment based on the Newcastle-Ottawa Scale (Wells et al., 2011; see Appendix C for the adapted version used in this study) for each included article. This tool is recommended for reviews of case-control studies. This identified any studies with a high risk of bias (Table 1) for exclusion in a sensitivity analysis.

**Table 1.** Table of Studies Included in the Systematic Review.

| **Study** | **N**<br>**(SZ)** | **N**<br>**(HC)** | **Task Domain** | **Task Details** | **Outcome** | **Prior** | **Prior Type** | **RoB** |
| --- | --- | --- | --- | --- | --- | --- | --- | --- |
| Abo Hamza et al. (2021) | 50 | 50 | Visual detection (pareidolia) | Report perceptions of faces in visual noise | No. illusory responses (seeing faces in pure noise) | Instructions: participants asked to detect faces | Cognitive | Low |
| Bansal et al. (2022) | 48 | 36 | Motion perception | Coherent motion direction of a random dot kinematogram changes half way through each trial. Participants report motion direction at the end of the trial. | Proportion of responses centred on initial motion direction | Motion direction at trial onset | Perceptual | Moderate |
| Bansal et al. (2022) | 42 | 34 | Motion perception | Replication sample with above task | Proportion of responses centred on initial motion direction | Motion direction at trial onset | Perceptual | Moderate |
| Cassidy et al. (2018) | 16 | 17 | Auditory (tone duration) perception | 'Context' sequence of tones (of varying duration) followed by a 'target' tone. Participants reproduce duration of target. | Assimilation bias: target duration biased towards context duration | Context sequence | Perceptual | Low |
| Catalano et al. (2020) | 37 | 29 | Visual detection | Report presence of target on left or right of screen | Reaction time; Accuracy | Non-social (arrow) or social (face with left-/rightward gaze) cues directing attention | Perceptual | Low |
| Chhabra et al. (2016) | 15 | 15 | Auditory (voice) detection | Report perceptions of voices (neutral words) in white noise | Response bias (for reporting voices) | Instructions: participants asked to detect voices | Cognitive | Moderate |
| Colbert et al. (2010) | 34 | 35 | Auditory (voice) detection | Report perceptions of voices (the word 'who') in white noise | Response bias (for reporting voices) | Instructions: participants asked to detect voices | Cognitive | High |
| Daalman et al. (2012) | 40 | 40 | Auditory (voice) detection | Hear sentences ending in noise or a word in noise. Word completions are either predicted or unpredicted based on (sentence) context. | No. top-down responses: hearing predicted word on unpredicted word or noise trials. | Sentence context creates prediction about word completion | Cognitive | Moderate |
| Fogelson et al. (2011) | 32 | 30 | Visual detection | 'Target' shape preceded by either a random or 'predictive' sequence (100% predictive of target). Participants asked to detect targets. | Reaction time | Sequence of shapes: participants told predictive sequence is 100% predictive of target | Cognitive | Low |
| Galdos et al. (2011) | 30 | 307 | Auditory (voice) detection | Trials feature white noise or white noise with either barely or clearly audible speech. Report perception of voice. | No. speech illusions (hearing speech in pure white noise) | Clearly audible speech trials create expectation of voices | Perceptual | Moderate |
| Gawęda & Moritz (2021) | 33 | 33 | Audiovisual (voice) detection | Report voices in non-verbal street noise | No. false perceptions (hearing words in subthreshold trials) | High expectancy trials also present written word and video of actor spelling word | Cognitive | Low |
| Gold et al. (2007) | 24 | 16 | Visual search | Search for target shape in array of distractors | Search efficiency: increase in reaction time as more distractors added | Efficient search on 'half-attended' trials required use of instructions to limit search to stimuli based on colour/shape | Cognitive | Moderate |
| Haarsma et al. (2020) | 30 | 32 | Audiovisual (phoneme) perception | Perceptual task: 'blend' of two phonemes feature alongside video of actor articulating one of the phonemes. Participants report dominant phoneme. Proportion of each phoneme varies until participants report maximal ambiguity (indifference point). | Change in indifference point compared to reference condition (no visual cue) | Video of actor articulating phoneme | Perceptual | Moderate |
| Haarsma et al. (2020) | 27 | 31 | Audiovisual (phoneme) perception | Cognitive task: same as perceptual task above except visual cue is phoneme written onscreen | Change in indifference point compared to reference condition (no visual cue) | Written phoneme: participants learn to associate this with the respective phoneme in training | Cognitive | Moderate |
| Horga et al. (2014) | 10 | 10 | Auditory (voice) detection | Report speech stimuli. Trials featured either non-speech, speech, or no stimuli. | Accuracy | Instructions: participants asked to detect voices | Cognitive | Low |
| Kowalski et al. (2024) | 116 | 51 | Audiovisual (voice) detection | Report voices in non-verbal street noise | No. false perceptions (hearing words in subthreshold trials) | High expectancy trials also present written word and | Perceptual (video) and cognitive | Low |

|  |  |  |  |  |  | video of actor<br>spelling word | (written<br>word) |  |
| --- | --- | --- | --- | --- | --- | --- | --- | --- |
| J. R.<br>Martin et<br>al. (2014) | 15 | 16 | Auditory<br>detection<br>(hysteresis):<br>ascending trials | Report whether stimulus<br>(scream/ spectrally-rotated<br>scream) present at end of the<br>trial. SNR increases during the<br>trial. | % reported signal at<br>the end of the trial | SNR at trial onset | Perceptual | Low |
| J. R.<br>Martin et<br>al. (2014) | 15 | 16 | Auditory<br>detection<br>(hysteresis):<br>descending trials | Same task as above except SNR<br>decreases during the trial. | % reported signal at<br>the end of the trial | SNR at trial onset | Perceptual | Low |
| B. Martin<br>et al.<br>(2017) | 27 | 23 | Visual detection | Respond to appearance of<br>target, which appears after short<br>or long delay. Temporal cue<br>predicts target onset, neutral cue<br>does not. | Reaction time:<br>temporal cue<br>speeds responses<br>for short delay trials | Temporal cue<br>predicts target onset<br>time | Cognitive | Low |
| Moritz &<br>Woodward<br>(2006) | 34 | 26 | Vision (figure<br>disambiguation) | Rate whether incomplete line<br>figure is accurately described by<br>a word. Figure is gradually<br>disambiguated. | Bias against<br>*disconfirmatory*<br>evidence:<br>difference in ratings<br>for an incorrect<br>hypothesis in<br>current versus<br>previous trial | Belief about contents<br>of figure on previous<br>trial | Cognitive | Low |
| Moritz & Woodward (2006) | 34 | 26 | Vision (figure disambiguation) | Same as above | Bias against *confirmatory* evidence: difference in ratings for a correct hypothesis in current versus previous trial | Belief about contents of figure on previous trial | Cognitive | Low |
| Norton & Chen (2011) | 24 | 21 | Motion perception | Report coherent motion direction of random dot kinematogram. Direction is either congruent or incongruent with previous trial. | Motion priming effect: difference in accuracy in congruent versus incongruent trials | Motion direction in previous trial | Perceptual | Moderate |
| Okruszek et al. (2018) | 46 | 40 | Visual (figure) detection | Point-light displays of two agents, A and B, engaged in communicative (e.g., 'squat down') or individual (e.g., 'sneeze') actions. Two intervals, only one containing B (masked in noise). Report which interval contains B. | Difference in $d'$ with communicative versus individual actions | Action by agent A: communicative actions facilitates detection of B. | Cognitive | Moderate |
| Okruszek et al. (2019) | 39 | 22 | Visual (figure) detection | Point-light displays of two agents, A and B, engaged in communicative or individual actions. B either replaced with or masked by noise. Report whether B is present. | Response bias: effect of action type on reports of B's presence | Action by agent A: communicative actions suggest B's presence. | Cognitive | Moderate |
| Pascucci et al. (2024) | 22 | 26 | Visual (orientation) perception | Report orientation of Gabor stimulus presented in peripheral visual field. | Serial dependence (score derived from modelling) | Orientation of Gabor on previous trial | Perceptual | Low |
| Pearl et al. (2009) | 14 | 10 | Audiovisual (phoneme) perception | One of two phonemes presented with a visual cue (actor producing articulatory movements for one of the phonemes). Report what phoneme is heard. | No. responses matching visual cue | Video of actor articulating phoneme | Perceptual | Moderate |
| Powers et al. (2017) | 29 | 15 | Audiovisual (tone detection) | Tone frequently presented with visual (checkerboard) stimulus (Pavlovian conditioning) in training. In testing, probability of tone presence decreases across blocks. Report whether tone is present on each trial | % conditioned hallucinations | (Implicit) learning of checkerboard–tone pairing | Perceptual | Low |
| Rohe et al. (2023) | 17 | 23 | Audiovisual detection | Sequences of simultaneously presented visual (flash) and auditory (beep) stimuli. Either congruent/incongruent numbers of flashes and beeps. Report number of beeps/flashes | $P_{\text{common}}$ (derived from modelling) indicates strength of causal prior | ‘Causal prior’: belief about the causal relationship between stimuli | Perceptual | Moderate |
| Rohe et al. (2023) | 17 | 23 | Audiovisual detection | Same as above | Variance of numeric prior (derived from modelling) indicates strength of numeric prior | ‘Numeric prior’: belief about the number of stimuli | Perceptual | Moderate |
| Romero et al. (2016) | 14 | 14 | Audiovisual (phoneme) perception | One of two phonemes presented with a visual cue (person making articulatory movements for one of the phonemes). Report the phoneme heard | % responses matching visual cue | Video of actor articulating phoneme | Perceptual | Low |
| Ross et al. (2007) | 18 | 18 | Audiovisual (word) perception | Hear word in noise alongside still image of face (auditory condition) or video of actor articulating the same word (audiovisual condition). Report the word heard | Difference in accuracy between auditory and audiovisual conditions | Video of actor articulating word | Perceptual | High |
| Schmack et al. (2015) | 29 | 32 | Motion perception | Intermittent presentation of stimulus (sphere formed from dots) with ambiguous rotation direction. Report direction of rotation. | Survival probability of percept across temporary stimulus removals | Reported motion direction before stimulus removal | Perceptual | Low |
| Schmack et al. (2017) | 19 | 25 | Motion perception | Presentation of stimulus (sphere formed from dots) with ambiguous rotation direction. Report direction of rotation. | Belief-induced bias ratio: ratio of belief-congruent and belief-incongruent mean percept duration | (False) information that glasses provided in task bias perceived direction of motion. Glasses paired with unambiguous stimuli in training to reinforce this belief | Cognitive | Low |
| Thakkar et al. (2021) | 36 | 22 | Visual perception | Negative Afterimage task: blurry disk (adaptor) with one bright half, one dark half followed by low contrast version of adaptor (nuller). Report which side of nuller is brightest. | Luminance contrast of nuller required to overcome aftereffect | Prolonged viewing of (adaptor) stimulus promotes illusion of 'photo-negative' afterimage | Perceptual | Low |
| Thakkar et al. (2021) | 36 | 22 | Visual (orientation) perception | Tilt Aftereffect task: flashing (adaptor) grating tilted 20 degrees. Nuller grating then presented at different angles. Report leftward/rightward orientation of nuller. | Tilt of nuller required to reach perception that grating is vertical | Prolonged viewing of (adaptor) stimulus promotes illusion of stimulus tilted in opposite direction | Perceptual | Low |
| Valton et al. (2019) | 20 | 23 | Motion perception | Field of moving dots presented at different contrasts. Report motion direction. | Estimation bias: difference between estimated mean and true motion direction | Statistical learning: two motion directions presented more frequently | Perceptual | Moderate |
| Vanes et al. (2016) | 40 | 22 | Audiovisual detection | Sequences of simultaneously presented visual (flash) and auditory (beep) stimuli. Either congruent/incongruent numbers of flashes and beeps. Report number of flashes. Fission condition: two beeps presented. | Difference in $d'$ (ability to distinguish between one and two flashes) for baseline (no beeps) versus fission condition | No. beeps | Perceptual | Low |
| Vanes et al. (2016) | 40 | 22 | Audiovisual detection | Same task as above but with fusion condition (one beep presented) | Difference in $d'$ (ability to distinguish between one and two flashes) for baseline (no beeps) versus fusion condition | No. beeps | Perceptual | Low |
| Van Leeuwen et al. (2021) | 18 | 23 | Visual detection | Letters, numbers, symbols embedded in noise (stimuli formed by illusory contours). Visibility of stimuli gradually increases (increasing phase) | Effect of decreasing phase prior. Perceptual gain (change in visibility threshold from | Prior created by viewing clearly visible stimuli at the end of the increasing phase | Perceptual | Moderate |
|  |  |  |  | then decreases (decreasing phase). Participants rate subjective visibility of stimuli at the end of each trial. | increasing to decreasing phase) |  |  |  |
| Van Leeuwen et al. (2021) | 18 | 23 | Visual detection | Same task as above but with synaesthesia-inducing stimuli | Effect of decreasing phase prior. Perceptual gain (change in visibility threshold from increasing to decreasing phase) | Prior created by viewing clearly visible stimuli at the end of the increasing phase | Perceptual | Moderate |
| Weintraub et al. (2012) | 21 | 22 | Auditory Segregation | Context sequence of tones with varying frequency separation precedes a (target) sequence with fixed frequency separation. Report whether target was perceived as a metronome (one stream) or galloping rhythm (two streams) | Proportion of trials with 'two-stream' reports | Context sequence: increasing frequency separation increases likelihood of perceiving 2 streams | Perceptual | Low |

*RoB– risk of bias; SZ– schizophrenia-spectrum psychosis patients; HC– healthy controls; N– number of participants*

### Meta-Analytic Modelling

Meta-analyses were conducted in RStudio (v4.2.2; R Core Team, 2022) using the metafor (Viechtbauer, 2010) and dmetar (Harrer, Cuijpers, Furukawa, & Ebert, 2019) packages. The dataset and analysis code are publicly available (<u>osf.io/sr39g</u>). The significance threshold for all analyses was 5%. All outcomes represented reliance on priors in patients versus controls to maximise their combinability. However, this meta-analysis pools effect sizes from different paradigms, hence heterogeneity was explored in further analyses, and the implications of this are discussed.

The first meta-analysis compared the reliance on priors in patients versus controls. Effect sizes for each experiment were either calculated using group means and standard deviations or extracted directly from the articles. To create a standardised scale for outcome measures, effect sizes were converted to the standardised mean difference Hedges’ (1981) *g*: a corrected version of Cohen’s *d* that has improved accuracy across sample sizes. Inverse values for *g* were computed where appropriate to ensure that positive values represented stronger priors in patients versus controls, while negative values indicated weaker priors in patients. These effect sizes were submitted to a random-effects, multi-level meta-analysis with studies weighted according to their precision. A random-effects model estimates the mean of a distribution of true effect sizes rather than a single true effect size (Hedges, Borenstein, Higgins, & Rothstein, 2009). This was appropriate due to the heterogeneity in study methods (different tasks and priors) and populations (different illness stages and diagnoses). A three-level meta-analysis was required to account for the nesting of experiments within studies. Some studies conducted multiple tasks or calculated effect sizes for different prior types with the same participants. Failing to account for this non-independence could generate false-positive results. The first level of the model represented participants (i.e., sampling variance of the effect sizes), who were nested within experiments (i.e., within-study variance across multiple tasks) at the second level, which were nested within studies (i.e., between-study variance) at the third level. This method enabled the inclusion of multiple results per study, avoiding the subjective exclusion of results. The model assumes independence of effect sizes between studies (at level 3), which is justified as there was no overlap of participant samples across studies.

A planned subgroup analysis was conducted to assess whether heterogeneity in effect sizes was due to different prior types. This used the same three-level model but included prior type as a dummy-coded fixed effect with two levels (perceptual versus cognitive priors). While this two-level model of priors is likely an oversimplification of a broader hierarchy, this enabled an initial investigation into factors that may explain conflicting results in a field that is yet to clearly define the predictive processing hierarchy. Research has proposed that this two-level model may reveal associations between psychosis and stronger priors at one level to compensate for weaker priors at another level (Haarsma et al., 2020). A meta-analysis implementing this theoretical framework appraises its plausibility and generalisability across tasks.

Two further meta-analyses investigated the correlation between prior strength and 1) hallucinations and 2) delusions in patients. Most studies reported correlation coefficients between measures of prior strength and symptom scores from various questionnaires. Pearson’s *r* values could therefore be extracted from these papers directly. A few studies instead featured effect sizes comparing reliance on priors in patient groups with and without the symptom, and therefore the effect sizes needed to be converted to Pearson’s *r*. All *r* values were then converted to Fisher’s *z* and submitted to a three-level random-effects model.

Three-level models estimate within-and between-study heterogeneity. *I^2^* was used as an overall estimate of whether heterogeneity significantly affected the analysis (Higgins & Thompson, 2002). Unlike Cochran’s (1954) *Q* on which it is based, *I^2^*is robust to the number of studies in the analysis. However, as *I^2^* represents the variability in effect sizes not due to sampling error, its value tends to 100% as the precision of studies increases. Thus, prediction intervals (PIs) were used to estimate the range of effect sizes that will likely be reported by future studies. PIs are an interpretable form of the heterogeneity statistic τ^2^ and are unaffected by the number of studies or their precision.

For all analyses, publication bias was assessed using funnel plots and Egger’s test. Any influential outliers, which were identified using forest plot visualisation and Cook’s distance, were excluded in a sensitivity analysis for each model.

## Results

### Search Results

Figure 1 presents the process of study selection. The search produced a total of 3,852 articles, of which 954 were duplicates. Title and abstract screening were therefore conducted on 2,898 articles, and 197 were identified as potentially relevant. Fifteen of these articles were not retrievable (e.g., poster presentations), thus 182 papers underwent full-text screening. Twenty-eight of these studies were combined with five studies identified through hand-searching, producing 33 studies to review. The dataset was comprised of 41 results as some studies featured multiple experiments. An overview of the included studies and demographics are presented in Tables 1 and 2, respectively.

**Figure 1.**
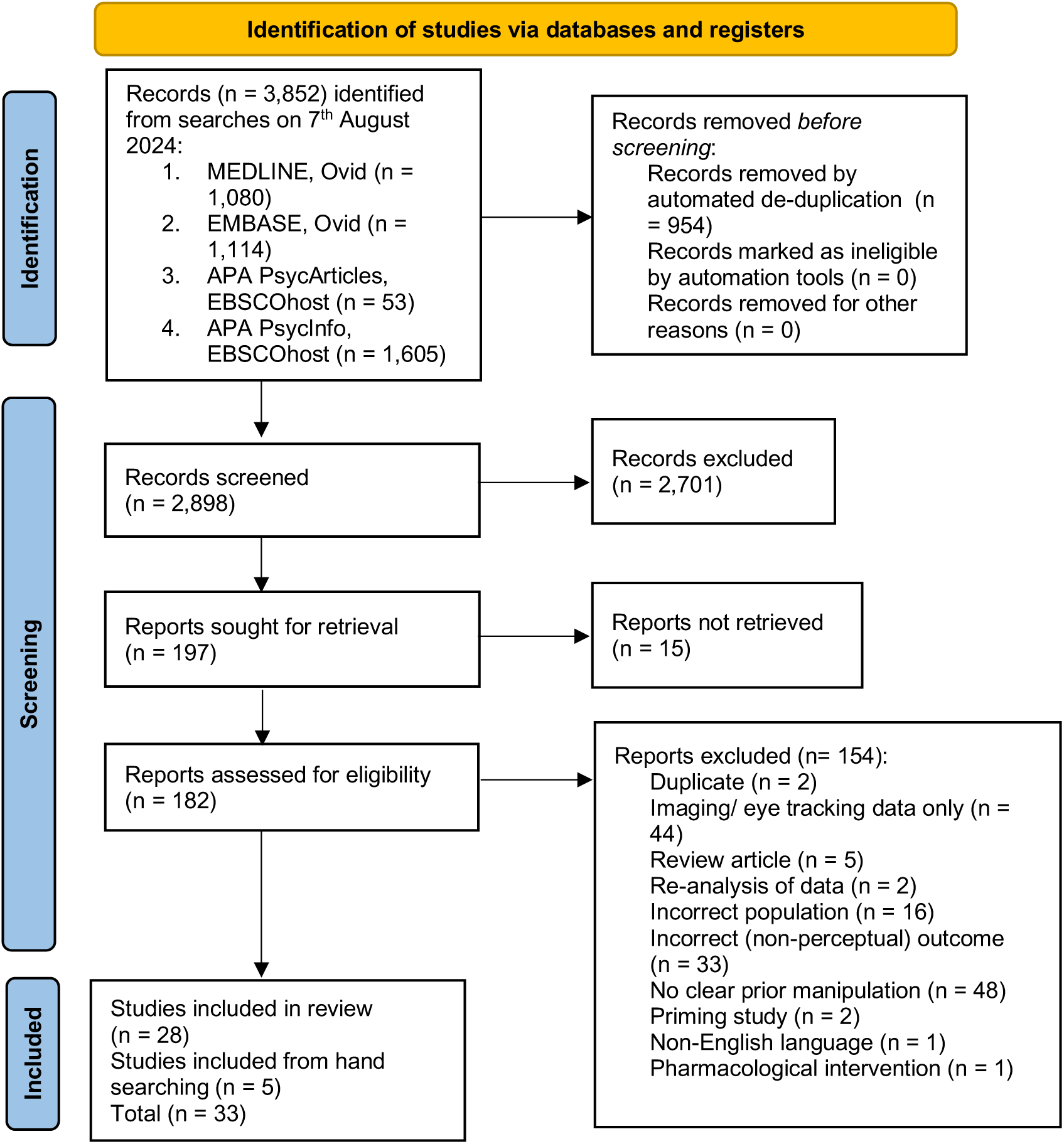
Preferred Reporting Items for Systematic Reviews (PRISMA; Page et al., 2021) Screening Procedure

**Table 2.**
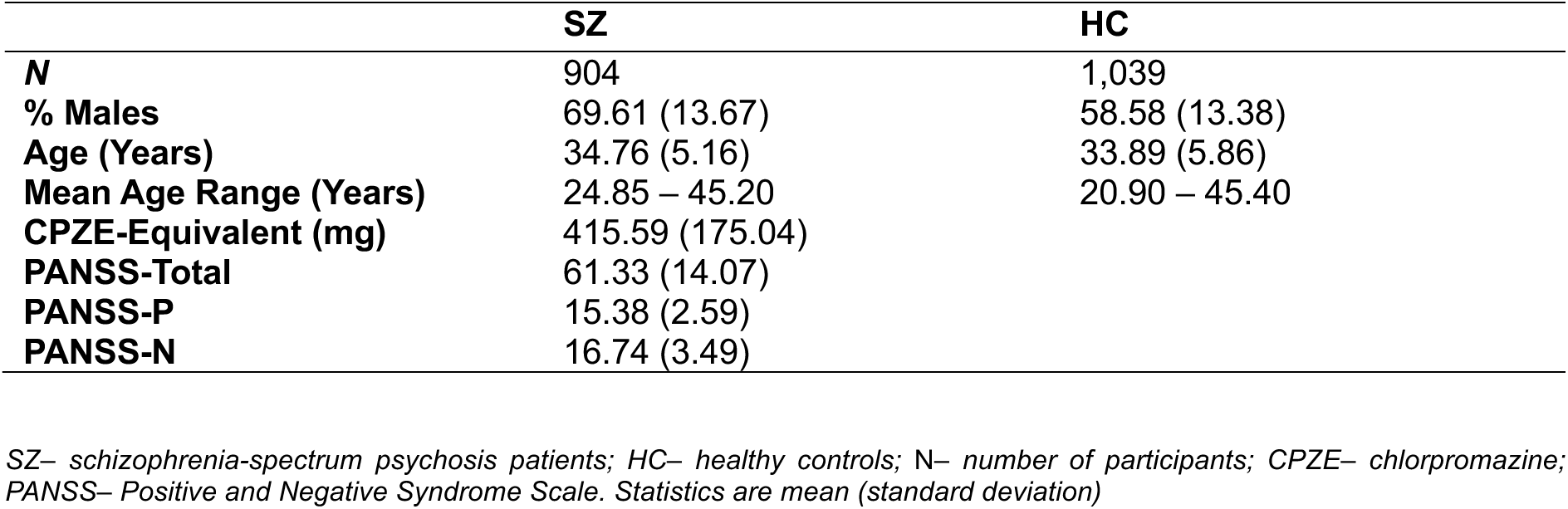
Demographics and Symptom Scores for Patients and Controls Across All Studies Included in the Meta-Analysis.

|  | **SZ** | **HC** |
| --- | --- | --- |
| **N** | 904 | 1,039 |
| **% Males** | 69.61 (13.67) | 58.58 (13.38) |
| **Age (Years)** | 34.76 (5.16) | 33.89 (5.86) |
| **Mean Age Range (Years)** | 24.85 – 45.20 | 20.90 – 45.40 |
| **CPZE-Equivalent (mg)** | 415.59 (175.04) |  |
| **PANSS-Total** | 61.33 (14.07) |  |
| **PANSS-P** | 15.38 (2.59) |  |
| **PANSS-N** | 16.74 (3.49) |  |

*SZ– schizophrenia-spectrum psychosis patients; HC– healthy controls; N– number of participants; CPZE– chlorpromazine; PANSS– Positive and Negative Syndrome Scale. Statistics are mean (standard deviation)*

### Relationship Between Illness Status and Prior Strength

Of the 41 results in the dataset, 34 results from 27 articles featured the information necessary to compute Hedges’ *g* (904 patients, 1,039 controls). Overall, there was no significant difference in the strength of priors in patients versus controls (*g* =.03, 95% CI [-0.27, 0.34]; *p* =.818). There was significant heterogeneity (*Q*(33) = 223.92, *p* <.001), the majority due to between-study heterogeneity (*I^2^ =* 70.37%). Non-significant between-group differences reported by individual studies were not due to the lack of a robust prior manipulation effect. The PI indicated that future studies could suggest either increased, decreased, or no difference in the reliance on priors in patients versus controls (95% PI [-1.51, 1.58]). A forest plot of the results is displayed in Figure 2.

**Figure 2.**
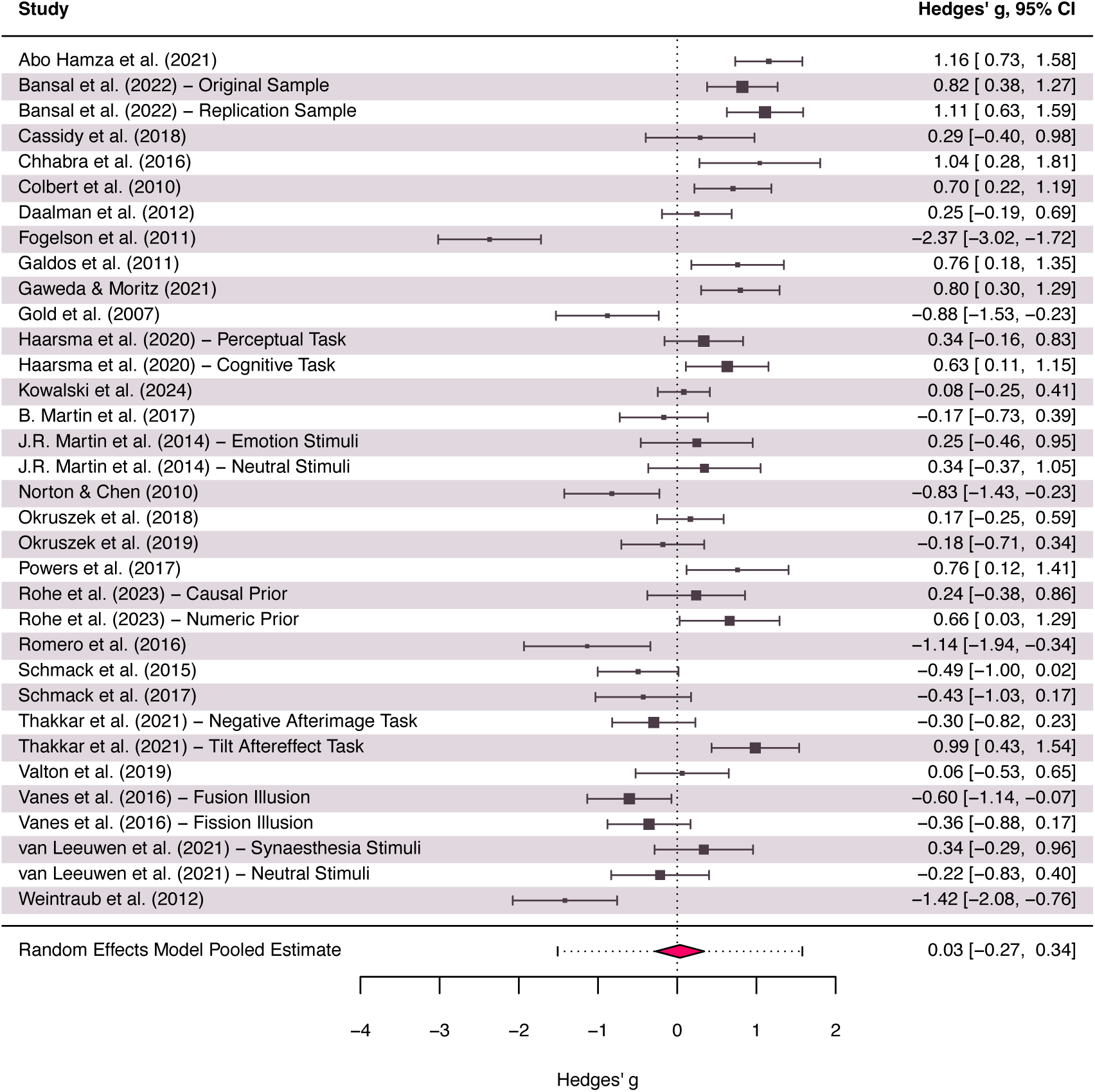
Forest Plot of Effect Sizes Representing Reliance on Priors in Patients Versus Healthy Controls Positive effect sizes represent stronger priors in patients compared to healthy controls. The vertical dotted line represents no difference in reliance on priors between groups. The true effect size (black rectangles) and 95% CI (confidence interval) are plotted for each included result. The size of the black rectangles is proportional to the weighting used to calculate the pooled effect size (red diamond). The red dotted error bar represents the 95% prediction interval. The width of the red diamond represents the 95% CI.

Sensitivity analyses were conducted to reduce the effect of bias on the results. Fogelson et al. (2011) was identified as an influential outlier, but the removal of this article did not significantly alter the pooled effect size. Another sensitivity analysis excluded Powers et al. (2017) given that additional comparisons by the authors demonstrated that stronger priors were associated with (clinical and non-clinical) hallucinations rather than psychosis. This finding was not accounted for in the meta-analysis, which focused on comparing patients and controls. Exclusion of this study did not significantly alter the results. Finally, excluding Colbert et al. (2010) due to a high risk of bias did not significantly change the results.

A funnel plot (Figure 3a) did not reveal evidence of significant publication bias and was corroborated by a non-significant Egger’s test intercept (*p* =.082; indicating no significant small study bias). The distribution of studies was mostly symmetric, with the exception of one outlier (Fogelson et al., 2011) that had already been addressed in sensitivity analyses. A wide distribution of effect sizes can be observed in the funnel plot, which likely reflects the heterogeneity of the studies included in the meta-analysis.

**Figure 3.**
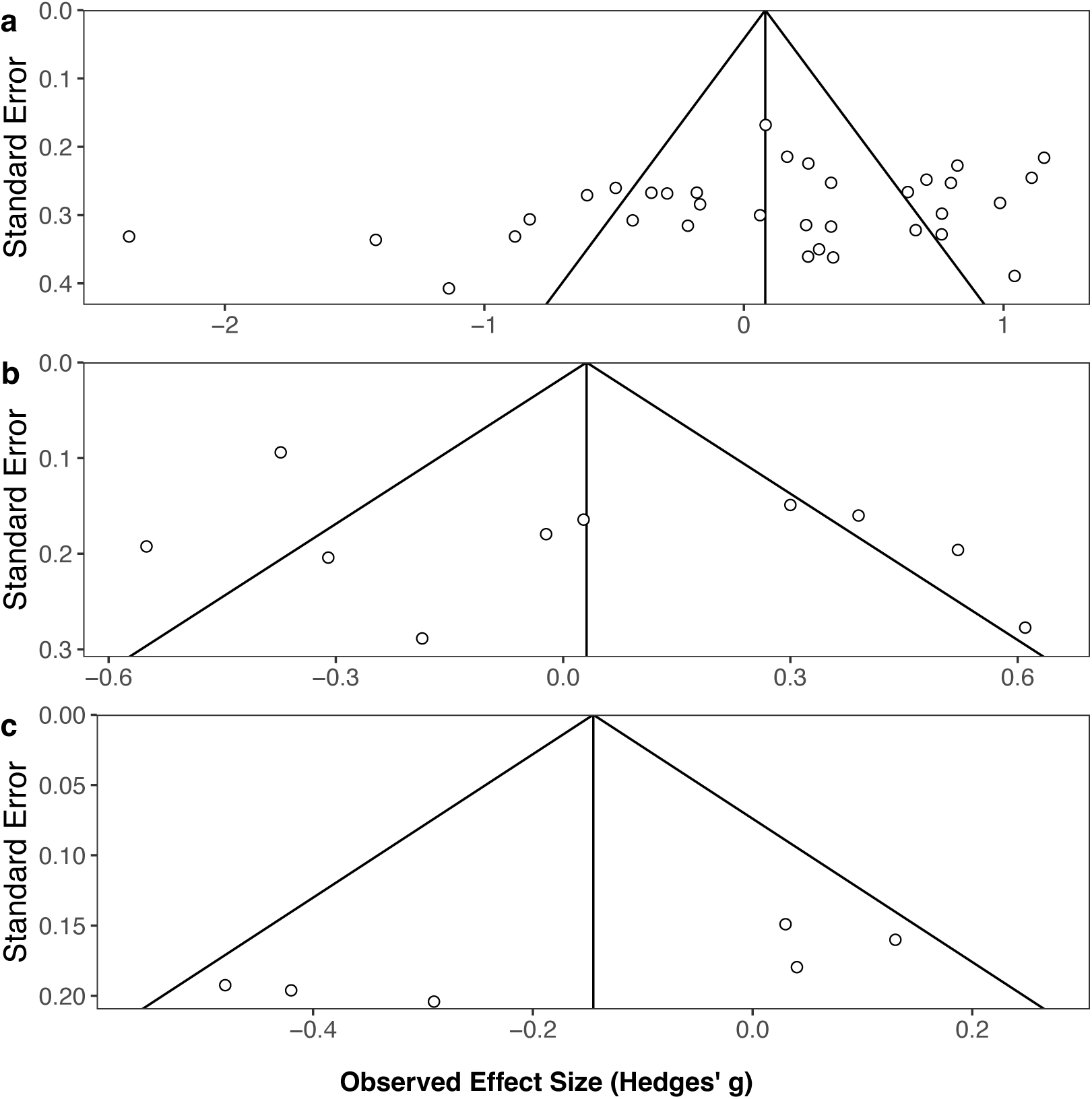
Funnel Plots of Study Precision (Standard Error) and Observed Effect Size. Plots represent analysis of **A**) reliance on priors in psychosis patients versus controls, **B**) correlation between reliance on priors and hallucinations in patients, and **C**) correlation between reliance on priors and delusions in patients.

The planned subgroup analysis provided an exploration of between-study heterogeneity. This included 21 perceptual and 13 cognitive prior effect results. There was no significant difference in the pooled effect sizes for the two prior types (*F*(1,32) = 0.1, *p* =.758), indicating that the two-prior model did not sufficiently explain differences in task results. Furthermore, neither reliance on perceptual priors (*g* =-.01, 95% CI [-0.41, 0.4]; *p* =.979) nor cognitive priors (*g* =.08, 95% CI [-0.48, 0.65; *p* =.758) significantly differed between patients and controls.

### Relationship Between Symptoms and Prior Strength

The meta-analysis focusing on the relationship between hallucinations and reliance on priors in patients incorporated 10 results from eight studies (370 patients; Figure 4). There was no significant relationship between hallucinations and prior strength (*r* =.04, 95% CI [-0.28, 0.36]; *p* =.780). There was significant heterogeneity (*Q*(9) = 47.81, *p* <.001). This was mostly between-study heterogeneity (*I^2^ =* 80.8%). The PI indicated that effect sizes for future studies could demonstrate a positive, negative, or no association between hallucinations and prior strength (95% PI [-0.81, 0.89]). A sensitivity analysis again excluded Colbert et al. (2010) due to a high risk of bias, but this did not significantly impact the results. A funnel plot (Figure 3b) revealed no evidence of publication bias, again supported by a non-significant Egger’s test intercept (*p* =.634). The spread of results indicated between-study heterogeneity.

**Figure 4.**
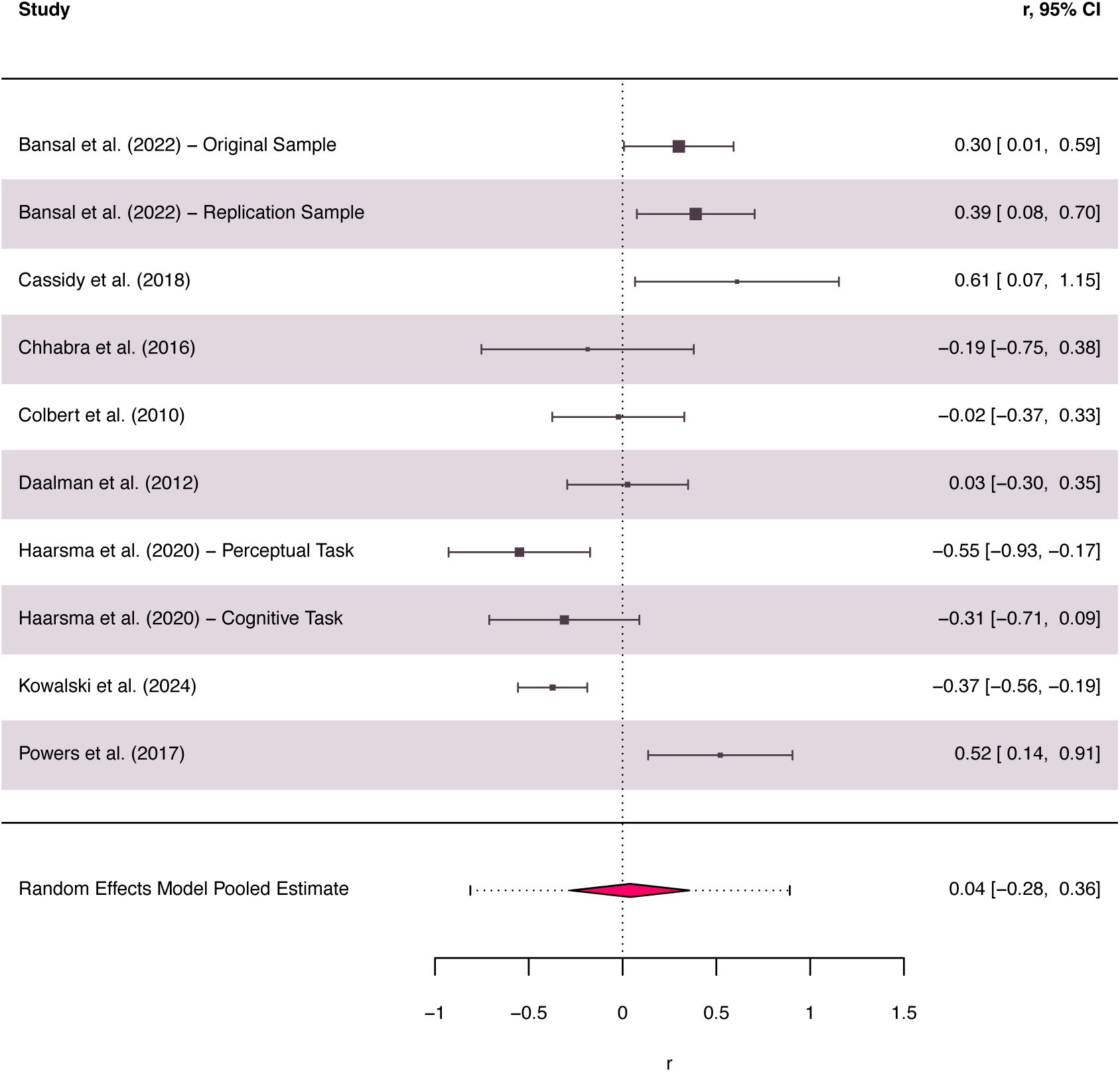
Forest Plot of Effect Sizes Representing Correlations Between Reliance on Priors and Hallucinations in Patients Positive effect sizes represent an association between stronger priors and hallucinations. The vertical dotted line represents no significant Pearson correlation (r) between reliance on priors and hallucinations. The true effect size (black rectangles) and 95% CI (confidence interval) are plotted for each included result. The size of the black rectangles is proportional to the weighting used to calculate the pooled effect size (red diamond). The red dotted error bar represents the 95% prediction interval. The width of the red diamond represents 95% CI.

The meta-analysis investigating the association between delusions and reliance on priors in patients incorporated six results from four studies (183 patients; Figure 5). There was no significant relationship between delusions and prior strength (*r* = -.16, 95% CI [-0.51, 0.19]; *p* =.293). There was significant heterogeneity (*Q*(5) = 10.78, *p* =.056), mostly due to between-study heterogeneity (*I^2^ =* 61.2%). The PI again indicated that effect sizes for future studies could demonstrate a positive, negative, or no association between delusions and prior strength (95% PI [-0.83, 0.51]). A sensitivity analysis again excluded Colbert et al. (2010) due to a high risk of bias, which did not significantly impact the results. A funnel plot (Figure 3c) did not clearly evidence publication bias, but interpretation was limited due to the small number of studies. The Egger’s test intercept did not indicate small study bias (*p* =.07).

**Figure 5.**
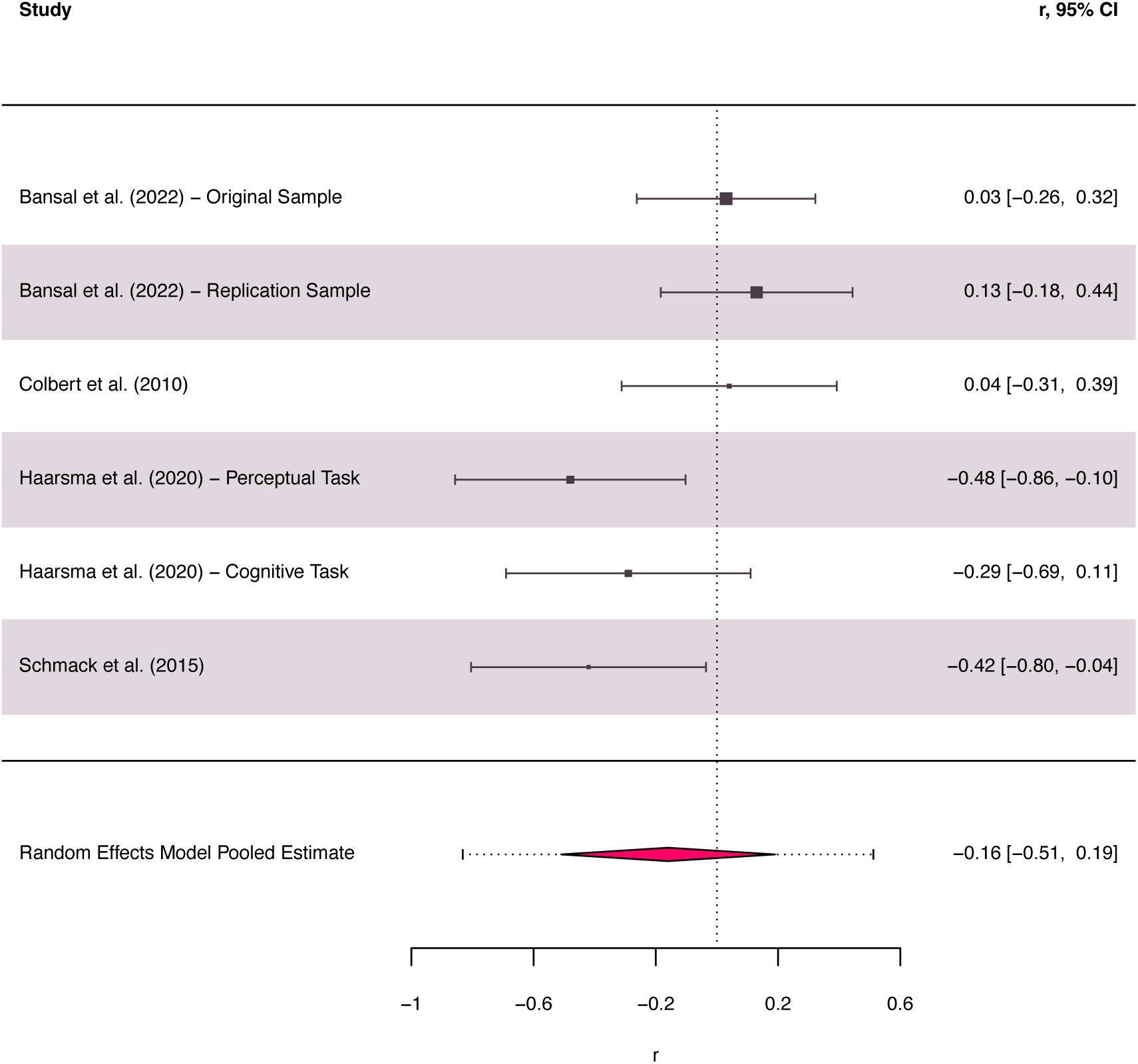
Forest Plot of Effect Sizes Representing Correlations Between Reliance on Priors and Delusions in Patients Positive effect sizes represent an association between stronger priors and delusions. The vertical dotted line represents no significant Pearson correlation (r) between reliance on priors and delusions. The true effect size (black rectangles) and 95% CI (confidence interval) are plotted for each included result. The size of the black rectangles is proportional to the weighting used to calculate the pooled effect size (red diamond). The red dotted error bar represents the 95% prediction interval. The width of the red diamond represents 95% CI.

Sub-group analyses also investigated the effect of prior type on the association between prior strength and symptoms. This is pertinent to the theory that an altered reliance on perceptual and cognitive priors may manifest as hallucinations and delusions, respectively. There was no significant moderating effect of prior type on the association between prior strength and delusions (*p* =.511; four perceptual, two cognitive tasks) or hallucinations (*p* =.188; five perceptual, five cognitive tasks) in patients. There was no significant correlation between hallucinations and reliance on perceptual priors (*g* =.2, 95% CI [-0.18, 0.58; *p* =.262), or delusions and reliance on cognitive priors (*g* = -.06, 95% CI [-0.63, 0.511; *p* =.790).

One limitation of the symptoms analyses was that the studies employed different symptoms scales. All studies except Colbert et al. (2010) used PDI scores for analyses of delusions. A previous sensitivity analysis already confirmed that excluding this study did not significantly affect the results. Measures of hallucinations were more variable and included comparisons of symptomatic and asymptomatic patients and scores on the PANSS-P3 (hallucinations subscale), CAPS, PSYRATS (Haddock, McCarron, Tarrier, & Faragher, 1999; Psychotic Symptom Rating Scales) auditory hallucinations subscale, and BPRS (Overall & Gorham, 1962; Brief Psychiatric Rating Scale). An exploratory meta-regression was therefore conducted to assess whether the difference in prior strength between patients and controls is significantly affected by patients’ mean PANSS-P (positive symptoms scale) scores. This analysis included 22 results from 17 studies (527 patients). There was no significant effect of PANSS-P scores on the difference in prior strength in patients versus controls (*F*(1,20) = 0.04, *p* =.84). This used a single scale to corroborate the results of the planned symptoms analyses, revealing no effect of positive symptoms on prior strength.

## Discussion

This study used a meta-analysis to systematically aggregate data from tasks investigating an altered reliance on priors in psychosis. Clarifying the role of predictive processing in psychosis is theoretically important and clinically relevant: it could inform the development of targeted cognitive interventions or digital phenotyping tools that monitor perceptual inference anomalies as early markers of relapse or treatment response. The high interest in this field is demonstrated by the large number of studies (*N* = 33) identified by our review, produced by multiple research groups across many countries. This literature features diverse tasks, reflected by the significant between-study heterogeneity. While a single pooled effect size is likely an inadequate summary metric for such heterogenous research, studies often extrapolate results across diverse tasks to advance a general theory of either stronger or weaker priors in psychosis. In contrast, the results of this study revealed no evidence for an altered reliance on priors in patients versus controls that can be consistently evidenced across paradigms. Importantly, this does not refute the possibility that aspects of aberrant predictive processing could contribute to mechanisms of psychosis. While the pooled effect size represented no psychosis-related alteration in reliance on priors, a number of individual studies reported significant between-group differences. For example, a subset of voice detection paradigms (Chhabra et al., 2016; Colbert et al., 2010; Galdos et al., 2011) indicated an increased reliance on priors in psychosis. However, even within this domain, associations with symptoms varied and therefore there was no clear relation to the clinical presentation of psychosis. Overall, the results indicate that psychosis is not associated with a generalised predictive processing alteration spanning multiple aspects of perception. Instead, predictive processing deficits in illness may be more nuanced. Our results recommend that authors exert caution when generalising conclusions across tasks and promote discussion of factors that may underlie conflicting results, with a view to creating a more refined cognitive model of illness.

As a comprehensive theory should explain common symptoms, further analyses examined whether altered prior strength accounts for hallucinations and delusions. The meta-analyses revealed no differences in prior strength related to either symptom, although these were based on a smaller subset of studies. A limitation of these analyses was that they combined correlations between reliance on priors and different symptom scales. Nevertheless, an exploratory meta-regression using PANSS scores also reported no effect of positive symptoms. A sub-group analysis also did not reveal evidence that distinct mechanistic pathways, namely reliance on perceptual and cognitive priors, underpin hallucinations and delusions, although this was again based on a small subset of tasks. This meta-analysis featured participants with both active and remitted symptoms, and there were insufficient studies to perform a sub-group analysis of the role of priors in each illness phase. Future work could address whether predictive processing deficits function as a state or trait marker of illness. These results could be useful for informing a predictive model of illness progression and for linking psychotic episode–related cognitive changes with underlying neurobiology and environmental risk factors (e.g., recent trauma). Future work should also explore previously-cited interactions between symptoms and illness stage (Haarsma et al., 2020) with a view to predicting transition to psychosis from the ARMS. Isolating possible medication effects on predictive processing could also inform pharmacocognitive models and drug selection, explain differences in task performance between patients, and help discern the effects of chronic illness and long-term medication exposure.

Possible reasons for heterogenous findings were explored in this review. Some theoretical arguments and individual studies (Haarsma et al., 2020; Schmack, Rothkirch, Priller, & Sterzer, 2017; Schmack et al., 2015) posit that conflicting results may be reconciled by considering a hierarchy of priors, broadly mirroring the predictive processing hierarchy (Hohwy, 2013). This has rarely been tested empirically and has not been investigated across paradigms with large samples. Our sub-group analysis indicated that between-study heterogeneity could not be resolved by differences in the use of perceptual and cognitive priors. This perceptual-cognitive distinction is frequently invoked in predictive processing literature (e.g., Haarsma et al., 2020), therefore our cross-study evaluation of this framework is theoretically relevant. However, this field lacks a clear delineation between these two prior types, with researchers often failing to reach a consensus on whether a given prior should be classified as perceptual or cognitive (Goodwin et al., 2026). This perhaps also reflects how a binary classification is likely a rudimentary model for describing a multi-level, graded hierarchy. Some of the included results also suggest that alternative, or additional, categories of priors could exist that may be differentially altered in psychosis. For example, Rohe, Hesse, Ehlis, & Noppeney (2023) used different classifications to evidence that between-group differences depend on prior type. Patients demonstrated an over-reliance on ‘numeric priors’ (beliefs about the number of stimuli) but were comparable to controls in terms of reliance on ‘causal priors’ (beliefs about the relationship between stimuli). This highlights how tasks may target distinct computational objects (e.g., sensory expectations, causal beliefs). Furthermore, two studies (Okruszek, Piejka, Wysokiński, Szczepocka, & Manera, 2018, 2019) reported patients display intact use of predictive cues (communicative gestures) for identifying biological motion relating to a second agent, despite lacking explicit recognition of these cues. Hence, impairments in predictive processing may depend upon the type of knowledge representation (i.e., explicit/implicit) required for the task. This finding highlights the possibility of alternative conceptualisations of the predictive processing hierarchy beyond the perceptual-cognitive framework. An implicit-explicit stratification could parallel the predictive processing hierarchy invoked in metacognition research, where the explicit evaluation of performance occurs as a second-order computation to performance (Fleming & Daw, 2017). Future work could explore a broader variety of prior types, including implicit priors developed across the lifespan, and psychosis-related alterations in Bayesian inference in cognitive and metacognitive tasks.

A challenge for predictive processing research is that different modelling frameworks can map parameters to priors in non-equivalent ways (Katthagen, Fromm, Wieland, & Schlagenhauf, 2022), potentially causing further discrepancies between study results. This review included outcomes based on various computational and behavioural metrics, which contributed to the heterogeneity of results. For example, Fogelson et al. (2011) reported prior-related between-group differences in reaction time, yet there was no effect of the prior on accuracy in either group. Analyses of multiple outcomes, or tracking how reliance on priors changes over time, could produce a more detailed evaluation than a single aggregate metric (Rohe et al., 2023; Weintraub et al., 2012). For example, the dynamic updating of priors could be particularly important for paradigms involving the tracking of statistical regularities or serial dependence (Weintraub et al., 2012).

Our findings align with the outcome of a systematic review on schizophrenia patients’ susceptibility to visual illusions (Costa et al., 2023). Although this previous study did not explicitly investigate the role of priors in illusory experiences, such phenomena are often deemed a product of top-down processing based on knowledge of natural scene statistics (de Lange, Heilbron, & Kok, 2018). Costa et al. (2023) did not indicate a general impairment for perceiving visual illusions in patients, and inconsistent findings were identified within each category of illusions. In place of a generalised deficit for perceiving illusions, the authors suggested that specific aspects of illusory tasks may be key to understanding anomalous processing in patients. Likewise, the present study observed conflicting results between tasks targeting the same cognitive domain (e.g., Daalman et al., 2012; Chhabra et al., 2016) and even replications of the same paradigm (Gawęda & Moritz, 2021; Kowalski et al., 2024). Further research is required to isolate task features that may potentially influence results and instigate differential processing in psychosis. One recurring theme identified by our review is the potential significance of uncertainty. Between-group differences may vary with sensory uncertainty and/or the precision of the prior. Cassidy et al. (2018) reported evidence that hallucination severity is associated with reduced uncertainty adjustment: the influence of context tones (the prior) on perception was not down-weighted as the variance of the tone sequence increased (i.e., as prior precision decreased). Sensory precision may also determine between-group differences. For example, Martin et al. (2014) identified a greater hysteresis effect (i.e., stronger priors) in patients but, for emotional stimuli, this was only observed with a decreased signal-to-noise ratio (i.e., lower sensory precision). Meanwhile, both Bansal et al. (2022) and Schmack et al. (2015) used random dot kinematograms to measure how motion perception is a function of previously encountered stimuli, yet these studies reported that psychosis is related to stronger and weaker priors, respectively. This discrepancy could be explained by differences in sensory uncertainty as Schmack et al. (2015) featured an ambiguous (bistable) stimulus, while Bansal et al. (2022) used a coherent motion signal embedded in noise. Between-group differences may also depend on how closely the task pertains to symptoms. Numerous tasks in this review measured the detection or hallucination of voices in noise following exposure to priors including explicit task instructions (e.g., suggestions of the presence of voices) and experience with clearly-audible voice trials. From these, a number of results indicated that patients over-weight priors compared to controls (Chhabra et al., 2016; Colbert et al., 2010; Galdos et al., 2011), indicating a possible target for future paradigms. It would also be important to investigate whether any psychosis-related expectation to perceive voices is causally related to illness or instead emerges following experience of symptoms (i.e., hallucinations).

While study selection for this review was performed against pre-registered inclusion criteria, conceptual difficulties concerning the definition of prior effects presented challenges. It remains possible that results included in the meta-analysis reflect mechanisms other than the use of priors. For example, patients’ altered audiovisual integration can be described by a preference for auditory stimuli (Pearl et al., 2009) rather than a dysfunctional weighting of priors. Moreover, there remain unresolved issues concerning how predictive processing operates, in particular the distinction between top-down and bottom-up processing. In this review, this was exemplified by contrasting effects observed in serial dependence tasks, where similar prior manipulations can induce either attractive effects (i.e., a bias towards perceiving the prior; Pascucci et al., 2024) or repulsive effects (i.e., a bias away from perceiving the prior, due to neuronal adaptation; Thakkar et al., 2021). One Bayesian account posits that repulsive effects reflect changes in the likelihood function (i.e., sensory information; Stocker & Simoncelli, 2005), which may be induced by prolonged exposure to high-contrast stimuli (Gekas & Mamassian, 2025). Relatedly, priming effects did not feature in this review given that these may be realised through bottom-up processing alone, although in some cases these may result from top-down or combined top-down and bottom-up processes (Rauss & Pourtois, 2013). Furthermore, Teufel & Fletcher (2020) discussed that priors may be instantiated through bottom-up and top-down mechanisms: predictions representing spatio-temporally global regularities (i.e., priors at the lowest level) may be coded by bottom-up processing, while top-down processing is reserved for context-dependent (higher-level) predictive signalling. Overall, clearer definitions of different types of priors and their relationship to top-down and bottom-up processing, as well as neurophysiological work linking this conceptual framework with functional activation and connectivity, would facilitate the design and interpretation of predictive processing paradigms and aid investigations into the mechanisms of psychosis.

In conclusion, this meta-analysis offers no evidence for a theory of altered prior strength in psychosis that can be generalised across perceptual tasks or mechanistically explains symptoms. Most studies only examined predictive processing from a narrow perspective (i.e., using a single task) yet many attempted to draw broad conclusions, and only one study included more than 50 patients. In future, the field would benefit from larger studies with multiple diverse paradigms conducted on the same patients, followed longitudinally across illness episodes where possible.

## Data Availability

All data produced in the present study are available upon reasonable request to the authors

## Appendix A: Search Strategy

### MEDLINE (via Ovid)

1. Psychos#s *.ti,ab,kw,kf*.
2. Psychotic *.ti,ab,kw,kf*.
3. Schizo* *.ti,ab,kw,kf*.
4. Schizophrenia/
5. Psychotic disorders/
6. 1 or 2 or 3 or 4 or 5
7. Belief* *.ti,ab,kw,kf*.
8. Believ* *.ti,ab,kw,kf*.
9. Prior* *.ti,ab,kw,kf*.
10. Expectation* *.ti,ab,kw,kf*.
11. Inference* *.ti,ab,kw,kf*.
12. “predictive processing” *.ti,ab,kw,kf*.
13. “predictive coding” *.ti,ab,kw,kf*.
14. “sensory attenuation”*.ti,ab,kw,kf*.
15. “sensory prediction*”*.ti,ab,kw,kf*.
16. 7 or 8 or 9 or 10 or 11 or 12 or 13 or 14 or 15
17. Percept* *.ti,ab,kw,kf*.
18. “sensory attenuation”*.ti,ab,kw,kf*.
19. “sensory prediction*”*.ti,ab,kw,kf*.
20. exp Perception/
21. orientation,spatial/
22. hearing/
23. smell/
24. taste/
25. touch/
26. vision, ocular/
27. thermosensing/
28. kinesthesis/
29. 17 or 18 or 19 or 20 or 21 or 22 or 23 or 24 or 25 or 26 or 27 or 28
30. 6 and 16 and 29
31. Limit 30 to yr=”2005-Current”
32. 31 not (exp animals/ not humans/.sh.)
33. Limit 32 to “review articles”
34. 32 not 33
35. limit 34 to english language

### EMBASE (via Ovid)

1. Psychos#s *.ti,ab*.
2. Psychotic *.ti,ab*.
3. Schizo* *.ti,ab*.
4. exp Schizophrenia spectrum disorder/
5. psychosis/
6. 1 or 2 or 3 or 4 or 5
7. Belief* *.ti,ab*.
8. Believ* *.ti,ab*.
9. Prior* *.ti,ab*.
10. Expectation* *.ti,ab*.
11. Inference* *.ti,ab*.
12. “predictive processing” *.ti,ab*.
13. “predictive coding” *.ti,ab*.
14. “sensory attentuation”*.ti,ab*.
15. “sensory prediction*”.*ti,ab*.
16. 7 or 8 or 9 or 10 or 11 or 12 or 13 or 14 or 15
17. Percept* *.ti,ab*.
18. “sensory attentuation”*.ti,ab*.
19. “sensory prediction*”.*ti,ab*.
20. exp Perception/
21. exp orientation/
22. smelling/
23. taste/
24. touch/
25. vision /
26. temperature sense/
27. “sensory dysfunction”/
28. 17 or 18 or 19 or 20 or 21 or 22 or 23 or 24 or 25 or 26 or 27
29. 6 and 16 and 28
30. Limit 29 to yr=”2005-Current”
31. Limit 30 to “remove medline records”
32. 31 not ((exp animal/ or exp invertebrate/ or nonhuman/ or animal experiment/ or animal tissue/ or animal model/ or exp plant/ or exp fungus/) not (exp human/ or human tissue/))
33. limit 32 to english language

### APA PsycInfo and APA PsycArticles (via EbscoHost)

1. TI psychos#s OR AB psychos#s
2. TI psychotic OR AB psychotic
3. TI schizo* OR AB schizo*
4. DE “Schizophrenia” OR DE “Acute Schizophrenia” OR DE “Catatonic Schizophrenia” OR DE “Childhood Onset Schizophrenia” OR DE “Paranoid Schizophrenia” OR DE “Process Schizophrenia” OR DE “Schizoaffective Disorder” OR DE “Schizophrenia (Disorganized Type)” OR DE “Schizophreniform Disorder” OR DE “Undifferentiated Schizophrenia”
5. DE “psychosis”
6. S1 OR S2 OR S3 OR S4 OR S5
7. TI belief* OR AB belief*
8. TI believ* OR AB believ*
9. TI prior* OR AB prior*
10. TI expectation* OR AB expectation* OR TI “sensory attenuation” OR AB “sensory attenuation” OR TI “sensory prediction*” OR AB “sensory prediction*”
11. TI inference* OR AB inference*
12. TI “predictive processing” OR AB “predictive processing”
13. TI “predictive coding” OR AB “predictive coding”
14. S7 OR S8 OR S9 OR S10 OR S11 OR S12 OR S13
15. S6 AND S14
16. TI percept* OR AB percept* OR TI “sensory attenuation” OR AB “sensory attenuation” OR TI “sensory prediction*” OR AB “sensory prediction*”
17. DE “Perception” OR DE “Anesthesia (Feeling)” OR DE “Apperception” OR DE “Auditory Perception” OR DE “Emotion Recognition” OR DE “Extrasensory Perception” OR DE “Form and Shape Perception” OR DE “Illusions (Perception)” OR DE “Intersensory Processes” OR DE “Mismatch Negativity” OR DE “Numerosity Perception” OR DE “Object Recognition” OR DE “Olfactory Perception” OR DE “Perceptual Closure” OR DE “Perceptual Constancy” OR DE “Perceptual Discrimination” OR DE “Perceptual Distortion” OR DE “Perceptual Learning” OR DE “Perceptual Localization” or DE “Perceptual Motor Processes” OR DE “Perceptual Organization” OR DE “Perceptual Orientation” OR DE “Perceptual Style” OR DE “Risk Perception” OR DE “Role Perception” OR DE “Self-Perception” OR DE “Sensory Gating” OR DE “Signal Detection (Perception)” OR DE “Social Perception” OR DE “Somesthetic Perception” OR DE “Spatial Perception” OR DE “Subliminal Perception” OR DE “Taste Perception” OR DE “Time Perception” OR DE “Visual Perception”
18. S16 OR S17
19. S15 AND S18
20. S15 AND S18 (narrow by population: animal)
21. S19 NOT S20
22. S21 (publication year 2005–2024) S22 (Narrow by Language-english)

### Appendix B: PRISMA 2020 Checklists

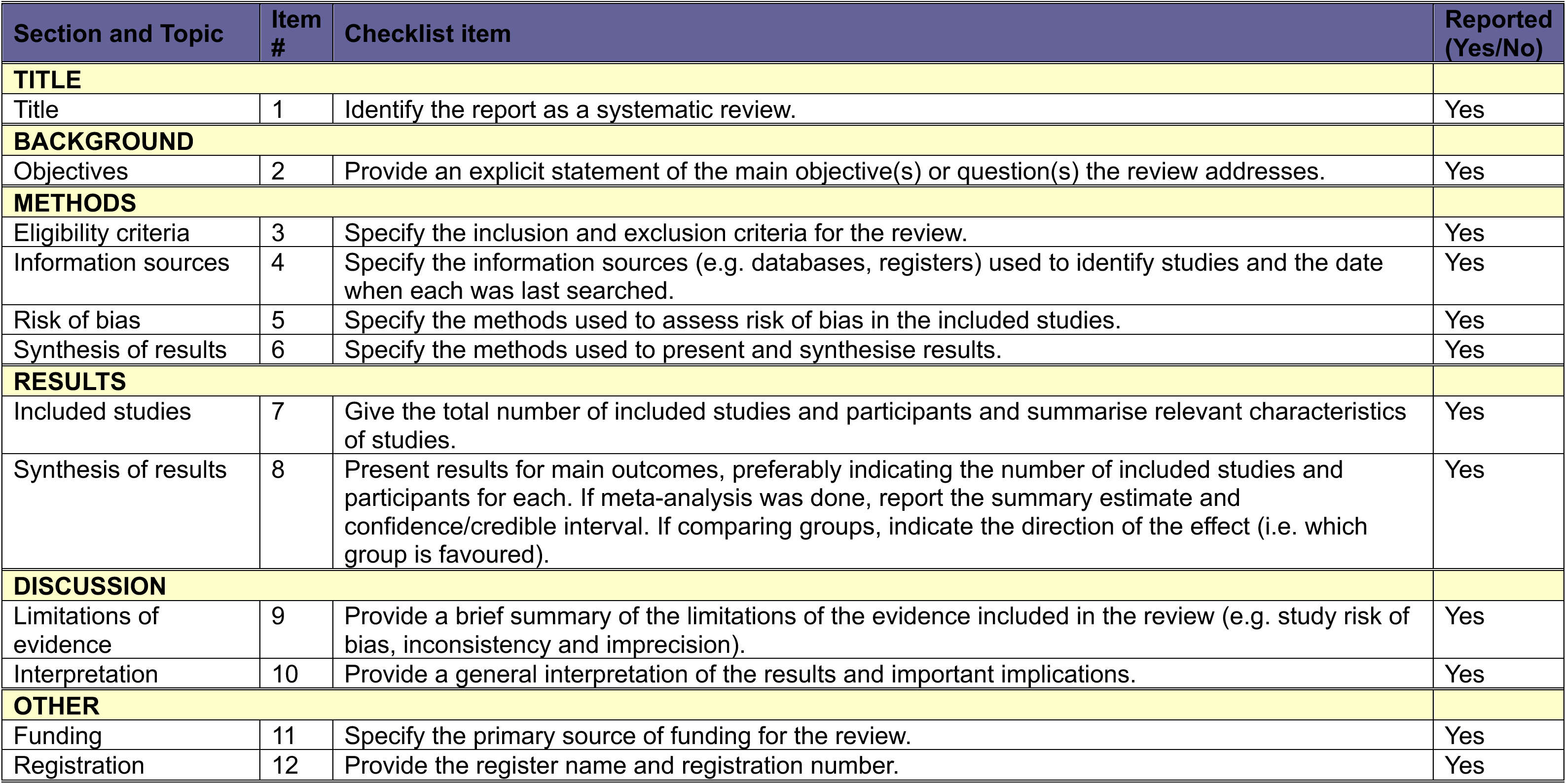
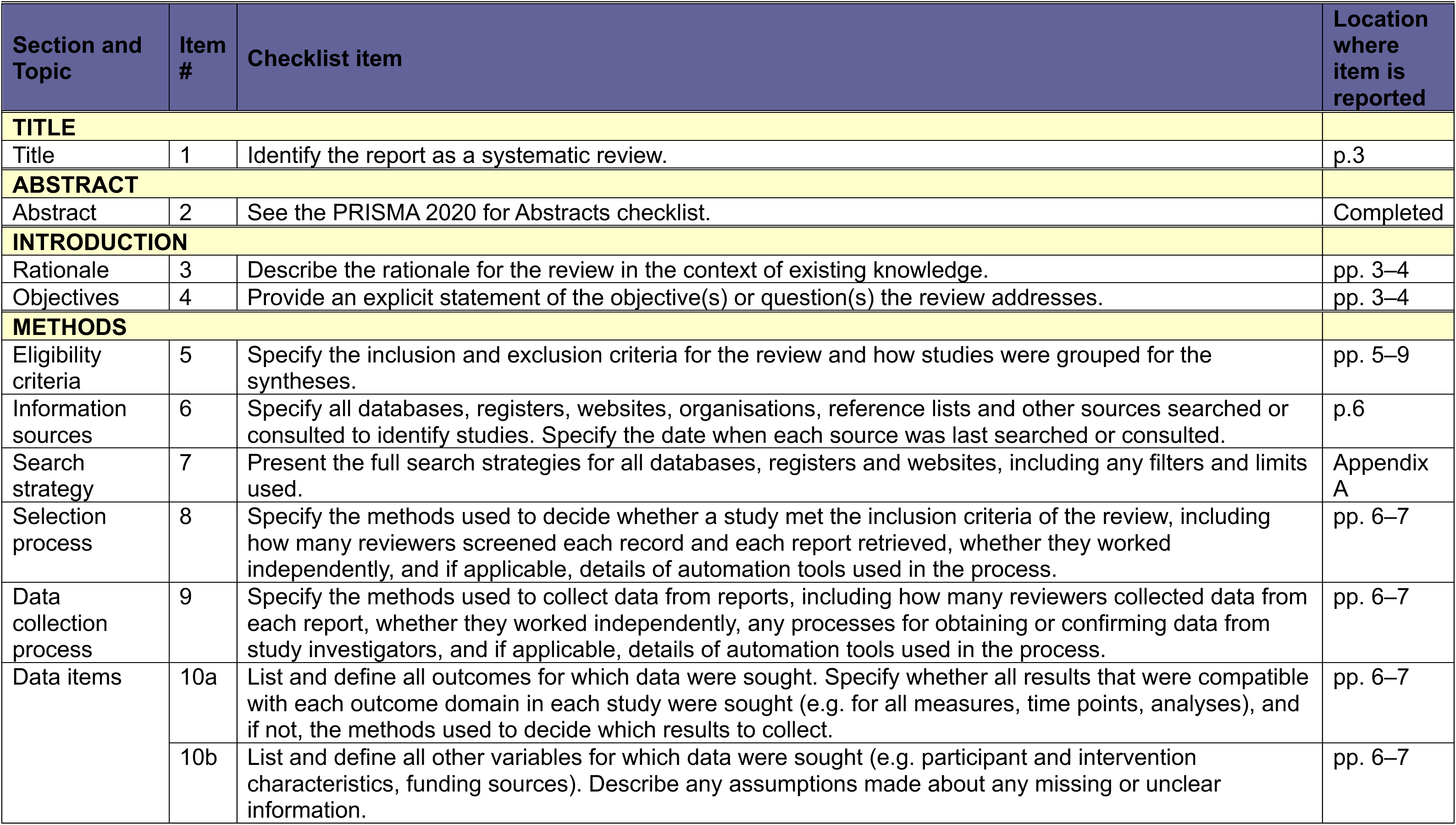
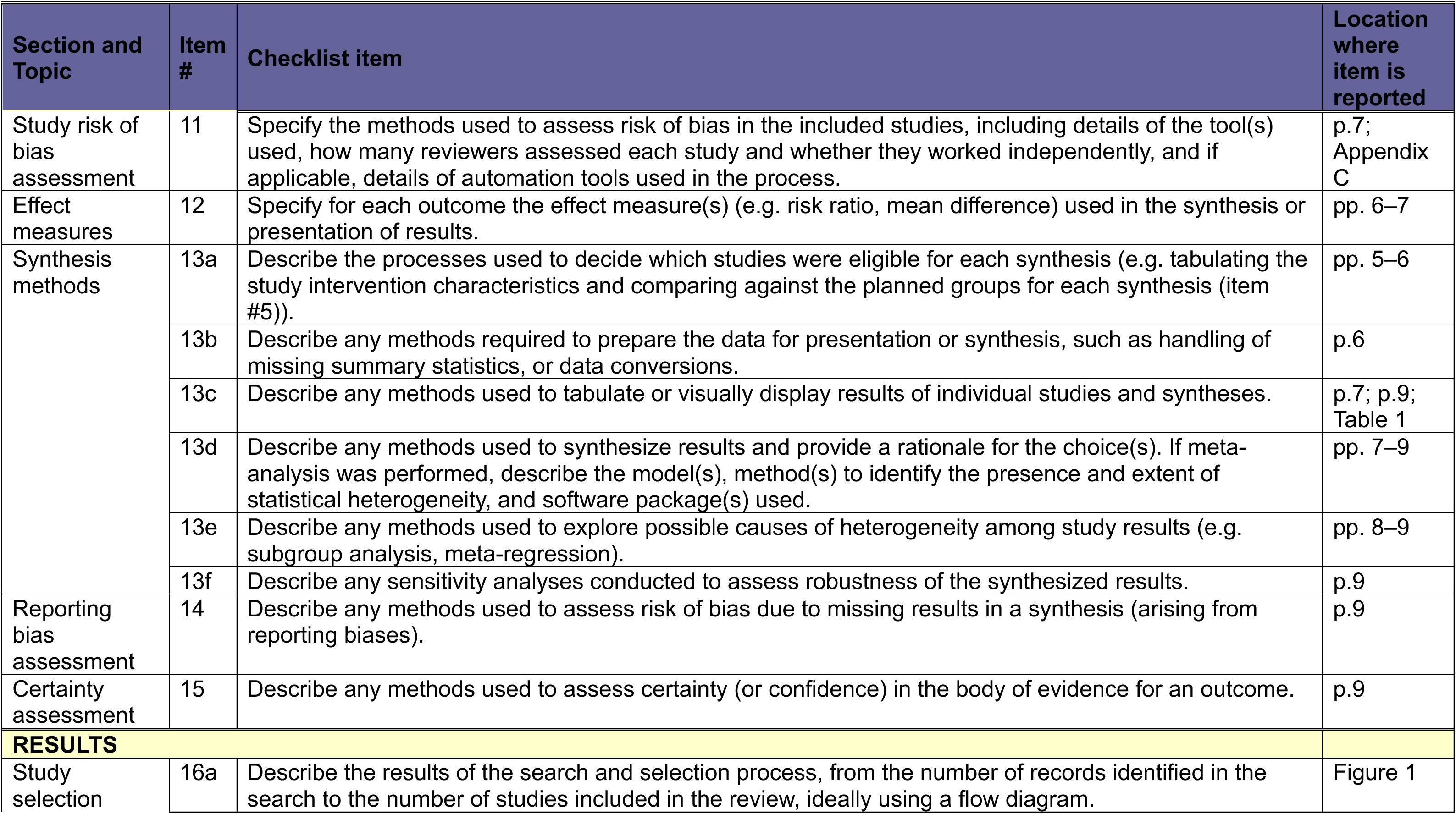
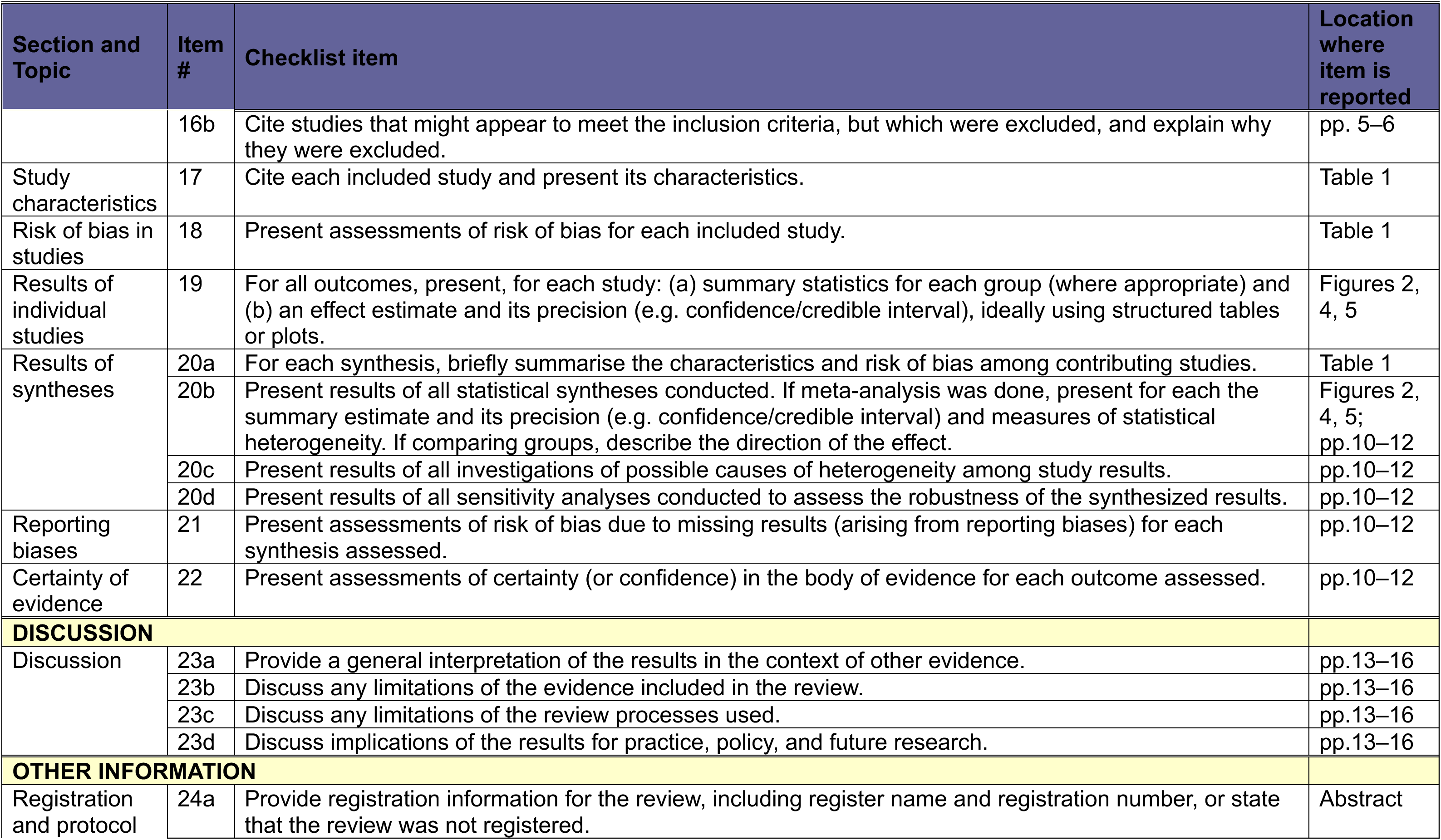
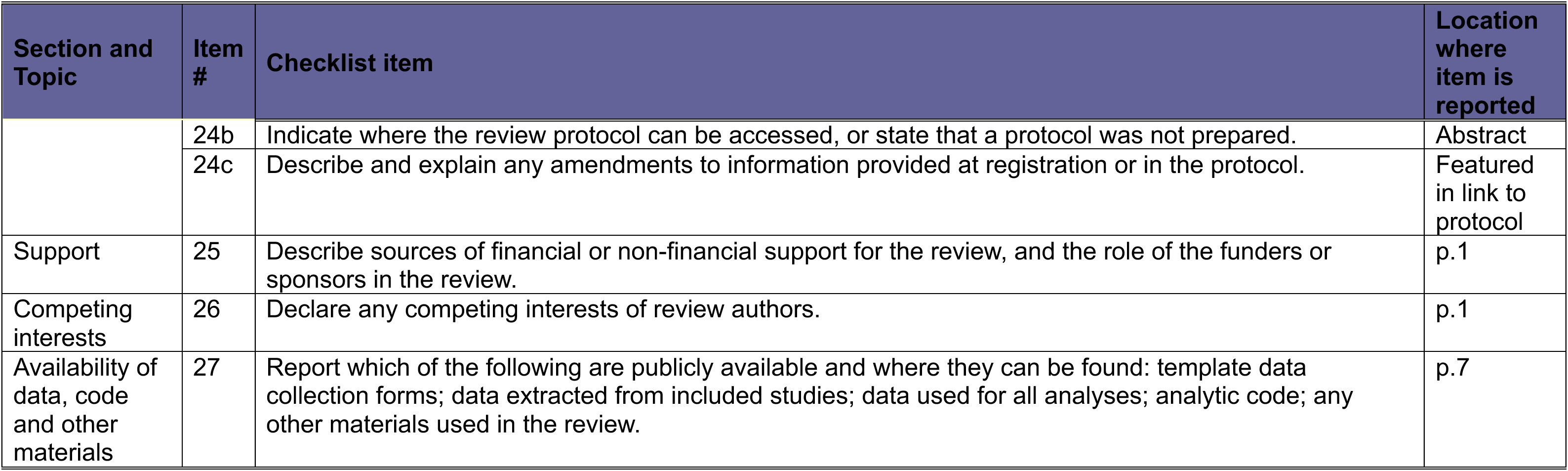

### Appendix C: Adapted Version of Newcastle-Ottawa Scale Selection

1. **Is there an adequate method used to identify patients?**

a. DSM, clinician/researcher diagnosis, medical records, etc *[1 point]*
b. Self-report, no description of diagnosis method included *[0 points]*
2. Were adequate exclusion criteria applied to patients?

a. No history of neurological disease or recent psychedelic drug misuse *[Must include both for 1 point]*
b. Either of the points in a) missing or not reported *[0 points]*
3. Is there adequate definition of control participants?

a. No history of psychiatric disease, neurological disorder, or recent psychedelic drug misuse *[Must include all 3 for 1 point]*
b. Any of the points in a) missing or not reported *[0 points]*

### Comparability

4. **Does either the study design (i.e., matching) or analyses control for confounding factors? Any covariates such as age, IQ, working memory, etc.**

a. 2 or more controlled for *[2 points]*
b. 1 controlled for *[1 point]*
c. No controlled for or not reported *[0 points]*

### Outcome

5. Does the task design allow a reliable assessment of reliance on priors?

a. Yes; the measure clearly reflects participants’ reliance on priors. There are different conditions to demonstrate the effect of priors. *[1 point]*
b. There are possible alternative explanations (not related to use of priors) that determine the outcome. [*0 points*]

## Notes

**Funding** C. M-S is funded by the Medical Research Council Doctoral Training Programme and Pinsent-Darwin Scholarship. This research received support from UKRI (MR/W020025/1). All research in the Department of Psychiatry, University of Cambridge is supported by the NIHR Cambridge Biomedical Research Centre (NIHR203312) and NIHR Applied Research Collaboration East of England. The views expressed are those of the author(s) and not necessarily those of the NIHR or the Department of Health and Social Care.

### Competing Interest Statement

The authors have declared no competing interest.

### Clinical Protocols

https://www.crd.york.ac.uk/PROSPERO/view/CRD42024574379

### Funding Statement

C. M-S is funded by the Medical Research Council Doctoral Training Programme and Pinsent-Darwin Scholarship. This research received support from UKRI (MR/W020025/1). All research in the Department of Psychiatry, University of Cambridge is supported by the NIHR Cambridge Biomedical Research Centre (NIHR203312) and NIHR Applied Research Collaboration East of England. The views expressed are those of the author(s) and not necessarily those of the NIHR or the Department of Health and Social Care.

### Author Declarations

The study used only openly available human data that were originally published in scientific journals.

